# Left atrial substrate characterization based on bipolar voltage electrograms acquired with multipolar, focal and mini-electrode catheters– the CHAZE-Substrate study

**DOI:** 10.1101/2023.01.24.23284964

**Authors:** Sven Knecht, Vincent Schlageter, Patrick Badertscher, Philipp Krisai, Florian Jousset, Florian Spies, Thomas Küffer, Antonio Madaffari, Beat Schaer, Stefan Osswald, Christian Sticherling, Michael Kühne

## Abstract

**Background:** Bipolar voltage (BV) electrograms for left atrial (LA) substrate characterization depend on catheter design and electrode configuration. The aim of the study was to investigate the relationship between the BV amplitude (BVA) using four different catheters and to identify their specific LA cutoffs for scar and healthy tissue.

**Methods:** Consecutive high-resolution electroanatomic mapping was performed using a multipolar Orion catheter (Orion-map), a duo-decapolar variable circular mapping catheter (Lasso-Map) and an irrigated focal ablation catheter with minielectrodes (Mifi-map). Virtual remapping using the Mifi-map was performed with a 4.5 mm tip-size electrode configuration (Nav-map). BVAs were compared in voxels of 3×3×3 mm^3^. The equivalent BVA cutoff for every catheter was calculated for established reference cutoff values of 0.1 mV, 0.2 mV, 0.5 mV, 1.0 mV, and 1.5 mV.

**Results:** We analyzed 25 patients (72% men, age 68±15 years). For scar tissue, a 0.5 mV cutoff using the Nav corresponds to a lower cutoff of 0.35 mV for the Orion and of 0.48 mV for the Lasso. Accordingly, a 0.2 mV cutoff corresponds to a cutoff of 0.09 mV for the Orion and of 0.14 mV for the Lasso. For a healthy tissue cutoff at 1.5 mV, a larger BVA cutoff for the small electrodes of the Orion and the Lasso was determined of 1.68 mV and 2.21 mV, respectively.

**Conclusions:** When measuring LA BVA in scar and healthy tissue, relevant differences were seen between focal, multielectrode and mini-electrode catheters. Adapted cutoffs for scar and healthy tissue are required.

## Introduction

Atrial fibrillation (AF) is associated with remodeling of the atrial myocardium.^1^ This substrate for AF initiation and perpetuation can be quantified using anatomical (echocardiographic, e.g. indexed left atrial (LA) volume (LAVI)), functional,^2^ structural (LGE MRI) or electrocardiographic measurement techniques for pre-procedural characterization. In order to demarcate diseased from healthy myocardium, invasive electrocardiographic measurements during the procedure are commonly performed using local bipolar voltage measurement from the electrodes of the catheters. These voltage maps may inform the physician on the presence or absence of LA substrate and may be used during substrate-based ablation strategies beyond pulmonary vein isolation. Since the bipolar voltage electrogram (BVA) reflects the local depolarization wavefront travelling along the electrode pair, it is strongly dependent on the configuration and orientation of the electrode pairs.

For the voltage characterization of the LA myocardium, several cutoffs have been proposed, ranging from 0.1 mV to 0.5 mV for the delineation of scar and between 0.5 mV and 1.5 mV for transition and healthy tissue (Supplemental Table). Validation of these cutoffs, however, is limited to a small sample size and initially was derived mainly from the thicker ventricular myocardium.^3,4^ With the advent of dedicated multipolar diagnostic catheter with different electrode configurations, however, these initially defined cutoffs determined with focal 3.5 mm- or 4 mm-tip ablation catheters were translated without validation and correlation between the measurements for the different devices. Nevertheless, these cutoffs are commonly used in clinical practice and have been applied to guide ablation. With a recent randomized-controlled trial showing clinical benefit of a substrate-based ablation strategy compared to PVI only, a standardized characterization and categorization independent of the type of catheter is urgently needed.^5^

The aim of the study was to compare BVA obtained from the LA using four different, established catheters and to deduce the individual, catheter-specific cutoffs for scar and healthy myocardial tissue.

## Methods

We prospectively included 25 consecutive patients referred for repeat catheter ablation for AF recurrence after index PVI. Only patients with recurrence after PVI were included in order to be able to analyse the full spectrum of electrical tissue characteristics in each patient, ranging from scar to healthy tissue. All patients signed informed consent prior to the study. The study (ClinicalTrials.gov Identifier: NCT04095559) was approved by the local ethics committee (Ethics Committee Northwest and Central Switzerland) and was conducted in accordance with the Declaration of Helsinki.

### Electroanatomical mapping

Consecutive electroanatomical mapping (EAM) using four different catheters was performed using an EAM system (Rhythmia, Boston Scientific, USA). In detail, ablation was performed under conscious sedation using fentanyl, propofol and midazolam. After venous access, a decapolar catheter (WEBSTER® CS Bi-Directional Catheter, Biosense Webster, USA) was inserted into the coronary sinus as an anatomical landmark for transseptal puncture and used as electrical reference for mapping. In patients in AF after access to the LA, electrical cardioversion was performed to restore sinus rhythm for subsequent substrate characterization. After detailed EAM using the multipolar Orion catheter (Intellamap Orion, 64 electrodes with 0.4 mm^2^ electrode size, 2.5 mm interelectrode spacing, Boston Scientific) (Orion-map), a remap on the same geometry was performed using a duo-decapolar variable circular mapping catheter (Lasso 2515 variable mapping catheter, electrode size 1 mm, interelectrode spacing 2-6-2 mm, Biosense Webster, Diamond Bar, USA) (Lasso-map) and an irrigated ablation catheter with minielectrodes (Intellatip Mifi OI, 4.5 mm tip electrode size, Mini electrode size 0.5 mm^2^ with 2.4 mm interelectrode spacing) (Mifi-map). A “virtual” remap of the Mifi map defining the Intellatip Mifi catheter as a focal 4.5 mm tip catheter without Mini electrodes (Intella Nav OI, ring electrode size 1 mm, interelectrode size of 2.5 mm) resulted in a fourth EAM of the LA (Nav-map). Bipolar voltage electrograms were filtered at 30-300 Hz.

### Data processing and analysis

The BVAs (peak-to-peak amplitude) in combination with the corresponding anatomical, three-dimensional location of the four electroanatomic maps of every patient were imported for data processing and analysis into Matlab (MATLAB. (2021). Natick, Massachusetts). Due to the skewed distribution of the voltage measures, a logarithmic transformation was applied to better fit a Gaussian distribution. Outlier voltages above 20 mV were excluded from the analysis.

Since point acquisition might have been inhomogeneous during mapping, resulting in unintended weighting of some regions, a voxel-based approach was performed to analyze and compare the maps and the corresponding BVA values. The mapping volume was divided in 3×3×3 mm^3^ cubes (voxels), and the mean of all mapping points within a voxel was calculated to characterize the myocardial tissue at this location (Figure 1).

**Figure 1:**
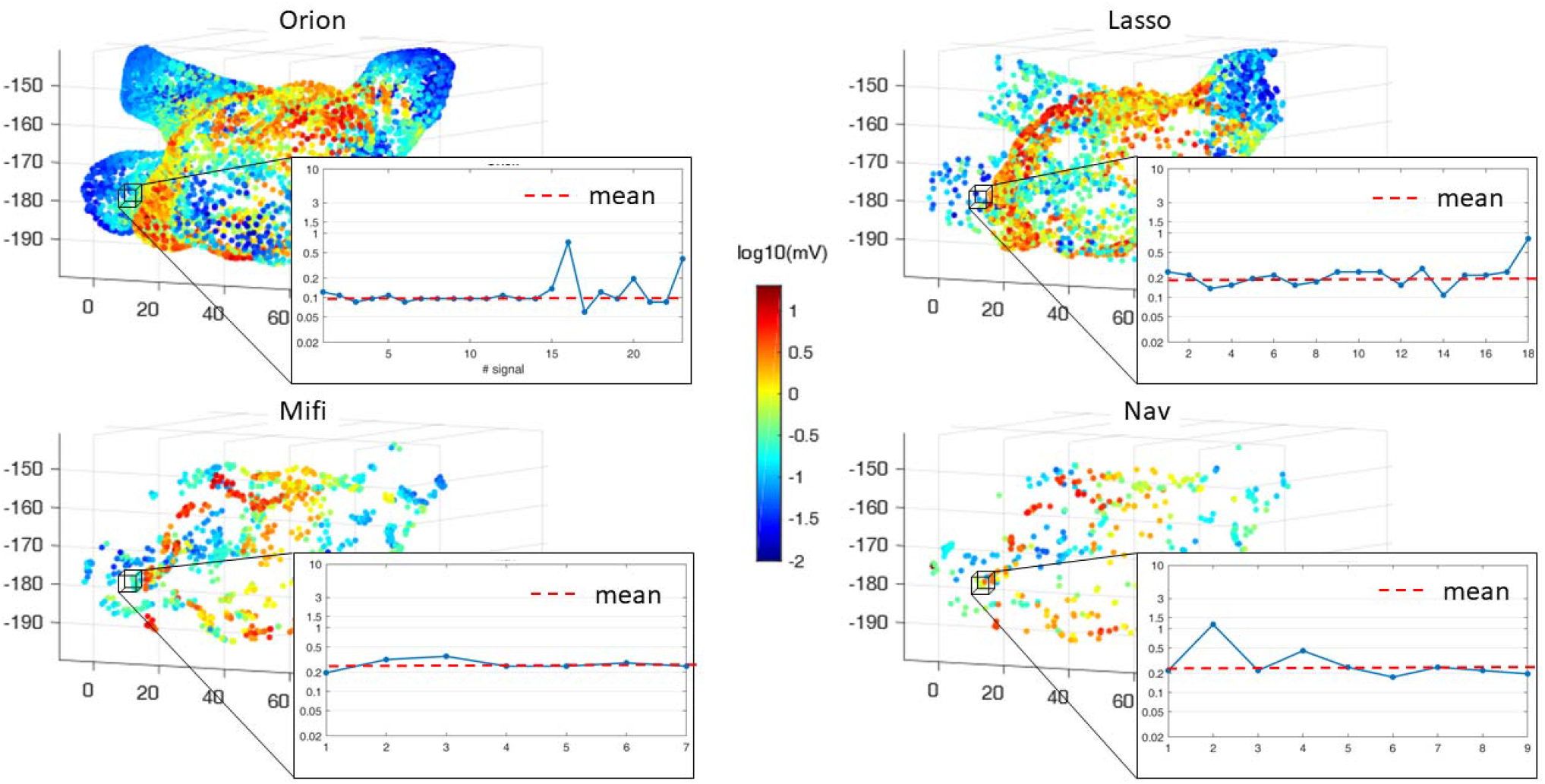
Exemplary representation of the BVA EAM using the Orion, Lasso, Mifi and Nav catheter. The black voxels represent the volume of 3×3×3 mm^2^ for which we calculated the mean voltage for catheter comparison (red dashed line) of all voltage values of this voxel.

Orthogonal regression (total least squares regression) was performed for every patient to investigate the relationship between the BVA measured by two different catheter types. The voltage pairs for the six possible combinations of the four catheter types (Orion-Lasso, Orion-Mifi, Orion-Nav, Lasso-Mifi, Lasso-Nav and Mifi-Nav) were analyzed for each patient. The goodness of fit of the regression is given by the ratio of the variance of the two principal components (ratio of the eigenvalues, 1 no correlation, 0 perfect linear correlation), and comparisons with low correlation (ratio >0.2) were excluded from analysis.

The multipolar Lasso catheter and the focal (Nav) catheter were used as references. To assess the matching between the cutoff values of these established catheter types and the novel catheter designs (IntellaMap Orion and IntellaNav Mifi OI), we used the following values of individual cutoffs for scar and low voltage classification of 0.1 mV, 0.2 mV, 0.5 mV, 1.0 mV, and 1.5 mV, respectively. For each cutoff, the mean equivalent cutoff over all patients was calculated. Values are given as mean ± standard deviation or median and interquartile range based on their distribution. Categorical data are shown as numbers and percentages.

## Results

We included 25 patients (72% men, age 68±15 years). Baseline data of the patients are summarized in Table 1. After EAM, repeat PVI alone was performed in 19 of the 25 patients (76%) and additional lesions including mitral isthmus line, roof line and cavotricuspid isthmus linear lesion were performed at the discretion of the treating physician in six patients (24%). The mean number of mapping points, the number of voxels and the number of paired voxels used for linear regression are summarized in Table 2.

**Table 1:** Baseline data of the patients.

|  | N=25 |
| --- | --- |
| Age [years] | 68±11 |
| Male sex | 18 (25%) |
| BMI [kg/m<sup>2</sup>] | 26±3 |
| Paroxysmal AF | 10 (40%) |
| Previous ablation |  |
| CB PVI | 7 (28%) |
| RF PVI | 18 (72%) |
| LVEF [%] | 59±8 |
| LA size (PLAX) [mm] | 41±8 |
| LAVI [ml/m<sup>2</sup>] | 38±9 |
| *Cardiovascular risk factors* |  |
| Hypertension | 16 (64%) |
| Dyslipidemia | 8 (32%) |
| CAD | 6 (24%) |
| Diabetes mellitus | 3 (12%) |
| Smoking |  |
| Never | 16 (64%) |
| Previous | 7 (28%) |
| Current | 2 (8%) |

BMI – body mass index, CAD – coronary artery disease, CB – cryoballoon, LA – left atrial,
LAVI – left atrial volume indexed, RF – radiofrequency.

**Table 2:**
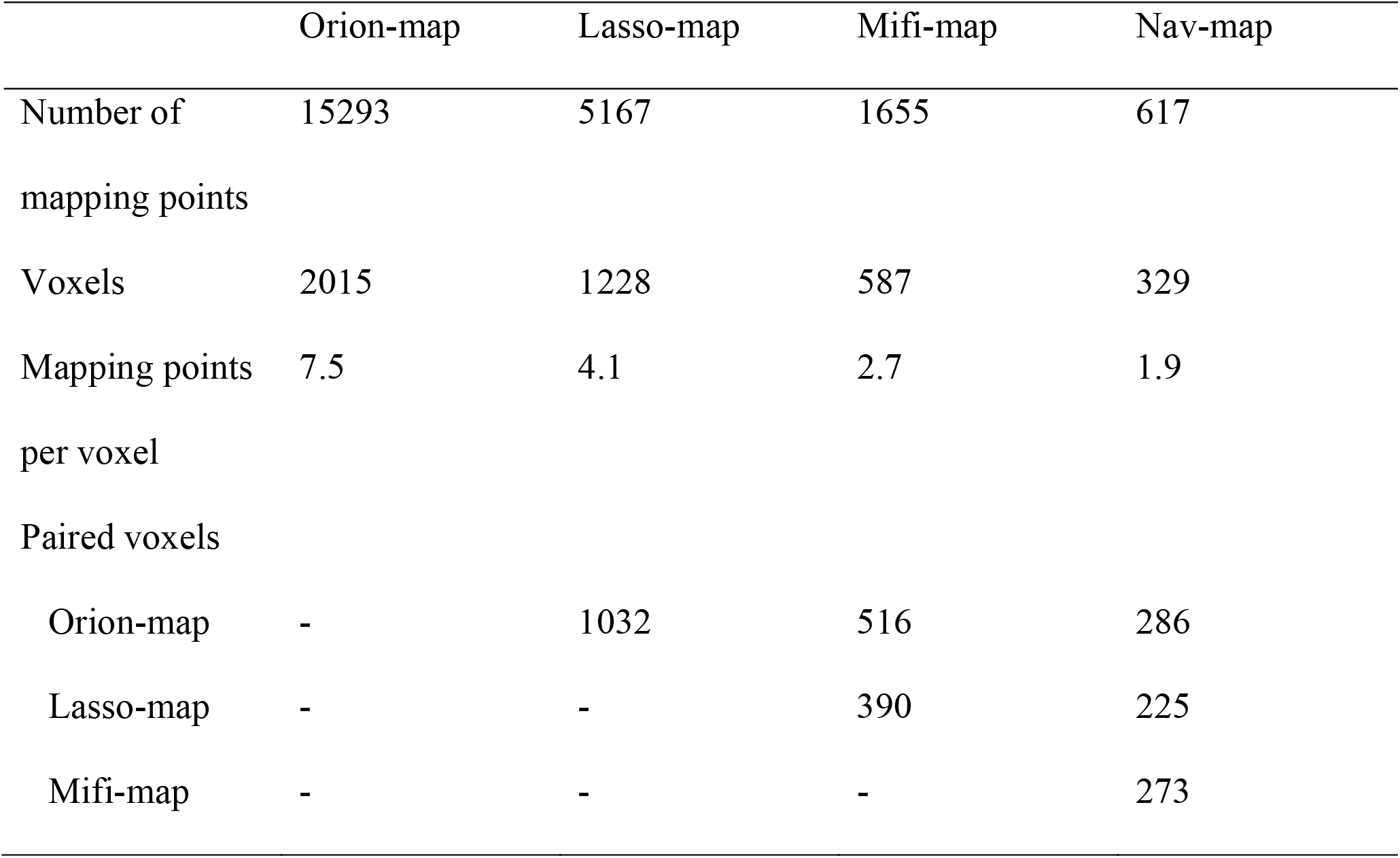
Characteristics of the electroanatomical maps

|  | Orion-map | Lasso-map | Mifi-map | Nav-map |
| --- | --- | --- | --- | --- |
| Number of mapping points | 15293 | 5167 | 1655 | 617 |
| Voxels | 2015 | 1228 | 587 | 329 |
| Mapping points per voxel | 7.5 | 4.1 | 2.7 | 1.9 |
| Paired voxels |  |  |  |  |
| Orion-map | - | 1032 | 516 | 286 |
| Lasso-map | - | - | 390 | 225 |
| Mifi-map | - | - | - | 273 |

### Correlation of the four catheters used for mapping

Correlation between the four different BVA maps for the 25 patients showed an individual variation between the 25 patients (Figure 2). This is as well visualized in the heat-map of the slope of the linear regression in Supplemental Figure 1. The mean slope was in the range of 56° for the Orion-Mifi relationship and 45° for the Mifi-Nav relationship. This 45° slope for the 4.5 mm tip (Nav) and the Mifi catheter results in a 1:1 association for all voltage values between these two electrode configurations.

**Figure 2.**
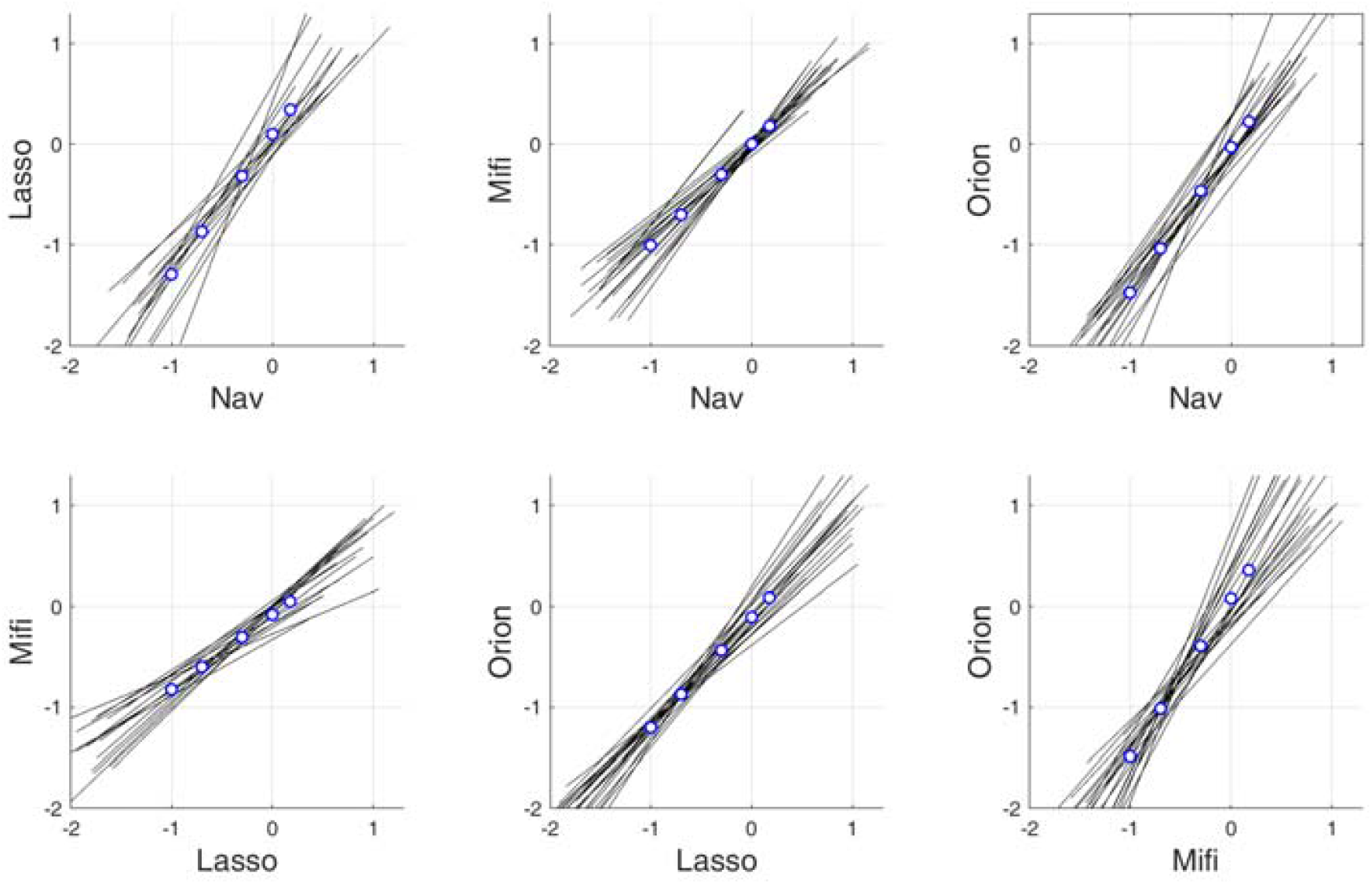
Orthogonal regression for all patients (black lines) in a logarithmic scale. Relations with low correlation (ratio >0.2) are excluded from analysis. The cutoff (mean value) for the catheter in the vertical axis, corresponding to a cutoff of 0.1 mV (−1), 0.2 mV (− 0.7), 0.5 mV (−0.3), 1.0 mV (0) and 1.5 mV (0.18) for the catheter in the horizontal, is calculated (blue circles).

### Cutoffs for scar and healthy tissue

The scar cutoff values of 0.1 mV, 0.2 mV, and 0.5 mV of the Nav catheter result in corresponding BVA cutoff values of 0.03 mV, 0.09 mV, and 0.35 mV for the Orion catheter, respectively. For the Lasso catheter the corresponding cutoff value was 0.05 mV, 0.14 mV, and 0.48 mV, respectively. When using Lasso catheter as comparator, the scar cutoff for the Orion catheter needs to be set at 0.06 mV, 0.13 mV, and 0.37 mV for the 0.1 mV, 0.2 mV and 0.5 mV cutoff value (Table 3).

**Table 3:** Summary of the individual, calculated voltage values for a specific catheter based on a specific reference catheter. The table links the voltage cutoff value used by the reference catheter to the corresponding catheter of interest.

|  | Reference value [mV] |  |  |  |  | Reference | Calculated<br>for |
| --- | --- | --- | --- | --- | --- | --- | --- |
|  | 0.10 | 0.20 | 0.50 | 1.00 | 1.50 |  |  |
| Calculated voltage values [mV] | 0.10 | 0.20 | 0.50 | 1.01 | 1.51 | Nav | Mifi |
|  | 0.05 | 0.14 | 0.48 | 1.26 | 2.21 | Nav | Lasso |
|  | 0.03 | 0.09 | 0.35 | 0.94 | 1.68 | Nav | Orion |
|  | 0.15 | 0.25 | 0.50 | 0.83 | 1.12 | Lasso | Mifi |
|  | 0.15 | 0.25 | 0.52 | 0.88 | 1.20 | Lasso | Nav |
|  | 0.06 | 0.13 | 0.37 | 0.79 | 1.22 | Lasso | Orion |
|  | 0.10 | 0.19 | 0.50 | 1.01 | 1.53 | Mifi | Nav |
|  | 0.05 | 0.14 | 0.52 | 1.39 | 2.47 | Mifi | Lasso |
|  | 0.03 | 0.10 | 0.41 | 1.20 | 2.27 | Mifi | Orion |
|  | 0.20 | 0.32 | 0.61 | 0.98 | 1.30 | Orion | Mifi |
|  | 0.21 | 0.34 | 0.65 | 1.08 | 1.44 | Orion | Nav |
|  | 0.15 | 0.29 | 0.69 | 1.31 | 1.91 | Orion | Lasso |

For healthy tissue with a 1.5 mV cutoff determined with the Nav catheter with a 4.5 mm-tip focal tip, the corresponding voltage cutoff for the Mifi, Lasso or Orion should be defined as 1.25 mV, 2.21 mV, and 1.68 mV, respectively. With the Lasso catheter as comparator, the cutoff of 1.5 mV corresponds to BVA value map in the range of 1.2 mV with the Mifi, Nav, Orion. (Table 3). A summary of the BVA cutoff values for an individual catheter type based on a reference map is shown in Table 3.

## Discussion

Bipolar voltage electrograms are strongly dependent on the catheter configuration defined by their electrode size and inter-electrode spacing. The relationship of the BVA between the various catheter designs with different electrode configurations is of high importance for the accurate and reproducible characterization and assessment of the LA substrate. The main findings of our study are: (1) Dependent on the catheter type used as a reference, a slight over- or underestimation of the BVA using the other catheter types was observed. (2) For areas with a low BVA, the Orion and the Lasso catheter showed a lower corresponding cutoff value for scar compared to the focal Nav catheter. Exemplarily, a 0.5 mV cutoff using the Nav corresponds to a cutoff of 0.35 mV for the Orion and of 0.48 mV for the Lasso, and a 0.2 mV cutoff corresponds to a cutoff of 0.09 mV for the Orion and of 0.14 mV for the Lasso. (3) For a healthy tissue cutoff above 1.5 mV based on the linear catheter, a larger BVA cutoff for the small electrodes of the Orion and the Lasso was determined of 1.68 mV and 2.21 mV, respectively.

### The impact of electrode configuration on the electrogram

To characterize the electric field generated by the cellular depolarization of the myocardium, electrodes are used for sensing and signal quantification. The measured unipolar voltage depends on the electric source and its temporal and spatial variation, which is determined by tissue characteristics, and on the electrode configuration. It is known that the amplitude of the unipolar voltage increases with a decrease in the electrode size (surface).^6,7^ As the surface area of the electrode has a spatial dimension, a certain percentage of the electrode surface might be further away from the electrical source. This results in a lower sensed “cumulative” voltage amplitude, since the voltage amplitude decreases with increasing distance from the electric source. This effect is especially important for bipolar voltage electrograms measured by large electrodes of a linear, focal ablation catheter like the herein used IntellaNav with a 4.5 mm tip size. A computational simulation for such a focal catheter showed that the BVA was significantly smaller for a catheter angulation of 45° with the ring electrode not being in contact with the tissue compared to a parallel orientation with both electrodes in contact.^8^ Furthermore, as shown in simulations and confirmed in an isolated porcine heart model, the BVA increases with increased interelectrode spacing up to a spacing of approximately 4 mm.^9^ In addition to the electrode configuration, the BVA is strongly influenced by the orientation of the activation wavefront relative to the electrode pair due to the signal cancellation of the unipolar voltage.^10^ This sensitivity on relative wavefront propagation on the BVA led to the development of direction-independent, multielectrode catheters and signal processing. ^11^

All these factors need to be considered when attempting to translate the voltage characteristics from one catheter type to another. We addressed these complex inter-relationships in our study by consecutive mapping of the same patient to define a universal table to link the established cutoff values for the different catheters. Furthermore, based on the voxel-based, averaged analysis, the impact of the orientation of the activation wavefront on the BVA might be reduced. We observed a linear relationship between the BVA values of the LA for all investigated catheter types over the entire voltage range of the LA. In contrast to the above described theory, however, we did not observe a larger BVA using the mini-electrodes of the Mifi catheter compared to the linear 4.5 mm tip-to-ring BVA. Another study with a different microelectrode catheter (Qdot, Biosense Webster, USA), however, showed larger BVA from the microelectrodes compared to the tip-to-ring BVA in the ventricle.^12^ This discrepancy might be explained by the even smaller electrode surface area and electrode distance of this catheter (0.167mm^2^ area and 1.755 mm interelectrode spacing) compared to the Mifi (0.4 mm^2^ and 2.4 mm interelectrode spacing). Furthermore, due to the proximal position of minielectrodes of the Mifi catheter (1.3 mm away from the tip), the electrogram is more sensitive to the catheter angulation, as described above. Finally, the behaviour of the (mini-) electrode size depends also on the electrical source. Whereas a significantly larger BVA of the minielectrodes was observed in healthy left ventricular myocardium, this difference diminished for low voltage scar tissue.^13^

### Voltage cutoff for scar and healthy tissue

The first bipolar voltage cutoffs for scar (<0.5 mV) and healthy myocardial tissue (>1.5 mV) were initially derived from ventricular myocardium using a 3.5 mm tip focal catheter.^3^ Despite the lower myocardial mass of the atrium compared to the ventricle, these values were translated to the atrium when using the same 3.5 mm tip irrigated ablation catheter. With the advent of multipolar catheters, such as the Lasso or the Orion catheter, the need for novel tissue-specific cutoffs emerged. Several cutoffs using different gold standards to define scar are currently available (Supplemental Table). In brief, currently available cutoffs are defined in the range between 0.1 mV and 0.5 mV for scar and 1.5 mV-2.2 mV for healthy tissue.

For the scar tissue with low voltage, we could show that the BVA is smaller for the Lasso and especially the Orion catheter compared to a 4.5 mm Nav catheter. Therefore, the scar cutoffs need to be set lower for these multipolar catheters. This difference is more pronounced with a lower reference cutoff: whilst the difference at a 0.5 mV cutoff is -30% for the Orion (0.35 mV compared to 0.5 mV), this increases to -70% with a scar cutoff of 0.1 mV using the Nav (0.03 mV compared to 0.1 mV). In conclusion, when using the same scar cutoffs, the low-voltage areas might be overestimated by the Orion (or Lasso) catheter (Supplemental Figure 2). This is in contrast to a comparison of a 3.5 mm tip irrigated focal catheter (Thermocool, Biosense Webster) with a pentaspline duo-decapolar catheter (Pentaray, Biosense Webster).^14^ Anter at al. showed that the low-voltage area (<0.5 mV) was larger for the linear catheter compared to the multipolar catheter. Since the pentaspline catheter has an identical electrode and size compared to the Lasso catheter, the difference might be most likely attributed to the differences of electrode size and spacing of the focal catheter. In contrast, in the ventricle, Berte et al. described larger low voltage areas (<0.5 mV) when mapping with the pentaspline catheter compared to the focal catheter.^15^ In a recent animal study on multipolar mapping catheters with the Pentaray (1 mm electrode size, 1 mm interelectrode spacing) compared to a novel Octaray catheter (Biosense Webster, USA) with smaller electrode size (0.5 mm electrode size, 1 mm interelectrode spacing) higher BVA amplitude in healthy as well as in previously ablated tissue was observed with the Pentaray catheter.^16^

This inconsistency among the published studies and the discrepancy from theory (according to which we would expect larger (unipolar) voltage amplitudes with smaller electrodes) confirms the necessity and relevance of our catheter-based instead of a purely theoretical assessment among the different catheter configurations.

## Limitation

First, this is a rather small study with 25 patients. However, a large number of points were acquired per map for intra-individual comparisons. Second, definite contact of the electrodes with the myocardial tissue could only be assured for the Nav and Mifi catheter with the force sensing technology, but even for these, the contact of the first ring-electrode with tissue could not be confirmed. This might have affected the measured amplitude of the BVA. Third, the local associations between the catheter types were performed on a voxel-based approach with a 3mm cubic voxel. This approach averaged potential deviations due to breathing or heart motion. Furthermore, the 4.5mm tip Intella Nav catheter with a different electrode size and configuration compared to the 3.5 mm tip ablation catheters from another manufacturer was used. Whether the same cutoffs hold true for these catheters is not clear. Finally, we did not test the duo-decapolar pentaspline catheter (Pentaray) in this study. Due to the same electrode size and configuration, consistent values compared to the Lasso catheter might be expected, but this was not shown.

## Conclusion

When measuring BVA in scar and healthy tissue of the LA, relevant differences were seen between focal, multielectrode and mini-electrode catheters used for mapping. Adapted cutoffs for scar and healthy tissue in the LA are required with different catheter configurations.

## Data Availability

Data available on request from the authors

## Disclosures

Sven Knecht has received funding of the “Stiftung für Herzschrittmacher und Elektrophysiologie” and the “Freiwillige Akademische Gesellschaft Basel”.

Patrick Badertscher has received research funding from the “University of Basel”, the “Stiftung für Herzschrittmacher und Elektrophysiologie” and the “Freiwillige Akademische Gesellschaft Basel”.

Beat Schaer reports speaker’s bureau for Medtronic.

Stefan Osswald received research grants from the Swiss National Science Foundation and Swiss Heart Foundation, Foundation for CardioVascular Research Basel, and F. Hoffmann-La Roche Ltd., and educational and speaker grants from F. Hoffmann-La Roche Ltd., Bayer, Novartis, Sanofi AstraZeneca, Daiichi-Sankyo, and Pfizer.

Christian Sticherling: Member of Medtronic Advisory Board Europe and Boston Scientitic Advisory Board Europe, received educational grants from Biosense Webster and Biotronik and a research grant from the European Union’s FP7 program and Biosense Webster and lecture and consulting fees from Abbott, Medtronic, Biosense-Webster, Boston Scientific, Microport, and Biotronik all outside the submitted work

Michael Kühne reports personal fees from Bayer, personal fees from Böhringer Ingelheim, personal fees from Pfizer BMS, personal fees from Daiichi Sankyo, personal fees from Medtronic, personal fees from Biotronik, personal fees from Boston Scientific, personal fees from Johnson&Johnson, grants from Bayer, grants from Pfizer, grants from Boston Scientific, grants from BMS, grants from Biotronik, all outside the submitted work.

Others have nothing to declare.

## Figures with Figure legends

**Central illustration:**
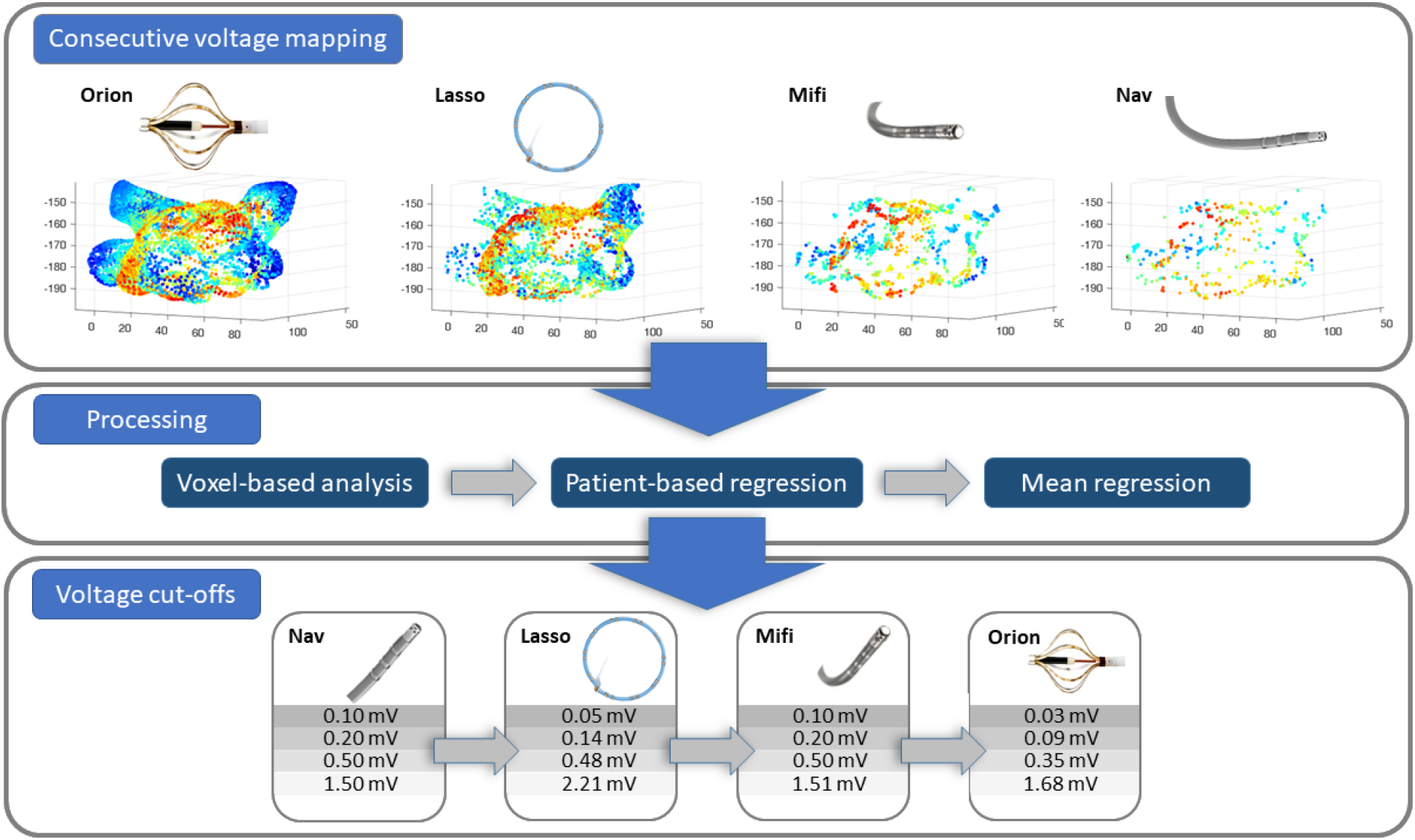
Pipeline of the analysis leading to the individual cutoff for every catheter based on the reference catheter value.

